# Policy and Effectiveness of Covid-19 Response

**DOI:** 10.1101/2021.05.27.21257908

**Authors:** Serge Dolgikh

## Abstract

A comparative analysis of selected national jurisdictions with respect to Covid-19 policy response indicated that factors such as effective communications and effective targeted intervention in the environments of higher epidemiological risk can be the key factors in the overall effectiveness of the epidemiological response minimizing both the impact of the epidemics and disruptions in the life of the society.

## 1 Introduction

Over the course of Covid-19 pandemic in 2020-21 different factors were analyzed as having a significant influence on the course of the epidemics [1-5]. At the time of writing it is expected that mass vaccinations being conducted in growing number of countries can become one of the key factors determining the development of the Covid-19 epidemiological situation going forward. At this point in time, before introduction of mass vaccination could significantly change the dynamics of the epidemics, it can be instrumental to examine how effective methods and approaches in the policy to control the spread of the epidemics and limit its impact on the society were among the group of national and subnational jurisdictions.

The aim of this work is to compare the policies in different jurisdictions selected by similarity with respect to a number of background factors, in two different avenues in the policy that emerged over this period: the quarantine-based policy including a number of factors associated with severity of its restrictions on the society; and the integrated management approach aimed at minimizing the epidemiological impact while avoiding strong disruptions to the life of the society. The principal method of the research is comparative statistical analysis by identified factors of interest as described in the following sections.

## 2 Methodology

A dataset of national and subnational epidemiological statistics as well as factors characterizing epidemiological policy and social factors at the time point of one year after the local arrival of the pandemics and before implementation of mass vaccination (pre-Vaccination Covid-19 (Northern) Epidemiology, pVCE) was used, compiled from sociological and epidemiological statistic obtained from open sources [6-10]. The impact of the epidemics was measured as reported mortality attributed to Covid-19, per 1 Million capita.

### 2.1 Selection Criteria

In selection of the cases (i.e., national and subnational jurisdictions, such as provinces) the following criteria were used to identify the cases with similar background factors and simply the analysis of significance of factors of interest:

- A minimal level of exposure to Covid-19 epidemics, identified as the minimum of 1,000 reported cases.
- A minimal population of 5 million, to reduce possibility of random fluctuations in the recorded parameters due to random nature of clusters of cases.
- Similar level of social development.
- Similar cultural and traditional background characterized as “northern European”.
- A minimal expectation of reliability and consistency of reported epidemiological information.
- Jurisdictions with high concentration of population, measured by 1) population density and 2) the number of municipalities (hubs) with population close to and over 1 Million, and a fraction of population residing in the hubs were excluded for consistency of the background as they would present substantially different social background for the analysis (Germany, France, possibly and/or in part, England, UK).

Applying these criterial ensured consistency of background factors among the cases in the dataset and allowed to concentrate on the factors of interest. Note that for large countries with highly non-homogeneous distribution of population as Canada and USA average national-level statistics cannot be considered very meaningful. These cases were not included in the statistical analysis and are shown only for completeness. A detailed analysis of policy and other factors in these cases on a subnational level could be a topic for another work.

Finally, it needs to be noted that any selection of data carries an inherent risk of introducing a bias. That being said, it is believed that the assumptions made and criteria used in the selection are reasonable from the perspective of identifying significant factors of influence; and based on these criteria the dataset is complete and prepared based on available data to the best of the author’s knowledge. Comments, observations, corrections are always welcome.

### 2.2 Dataset

The resulting dataset of national and subnational jurisdictions described by policy and social factors is presented in Tables 1,2. Policy factors included: public communications; severity of quarantine policy; engagement and voluntary participation of population; effectiveness of targeted intervention; general state of public healthcare system (PHC); epidemiological preparedness of PHC and quality of the management of epidemiological aspects by PHC, including resourcing, capacity, epidemiological safety; availability of specific effective treatments.

**Table 1.**
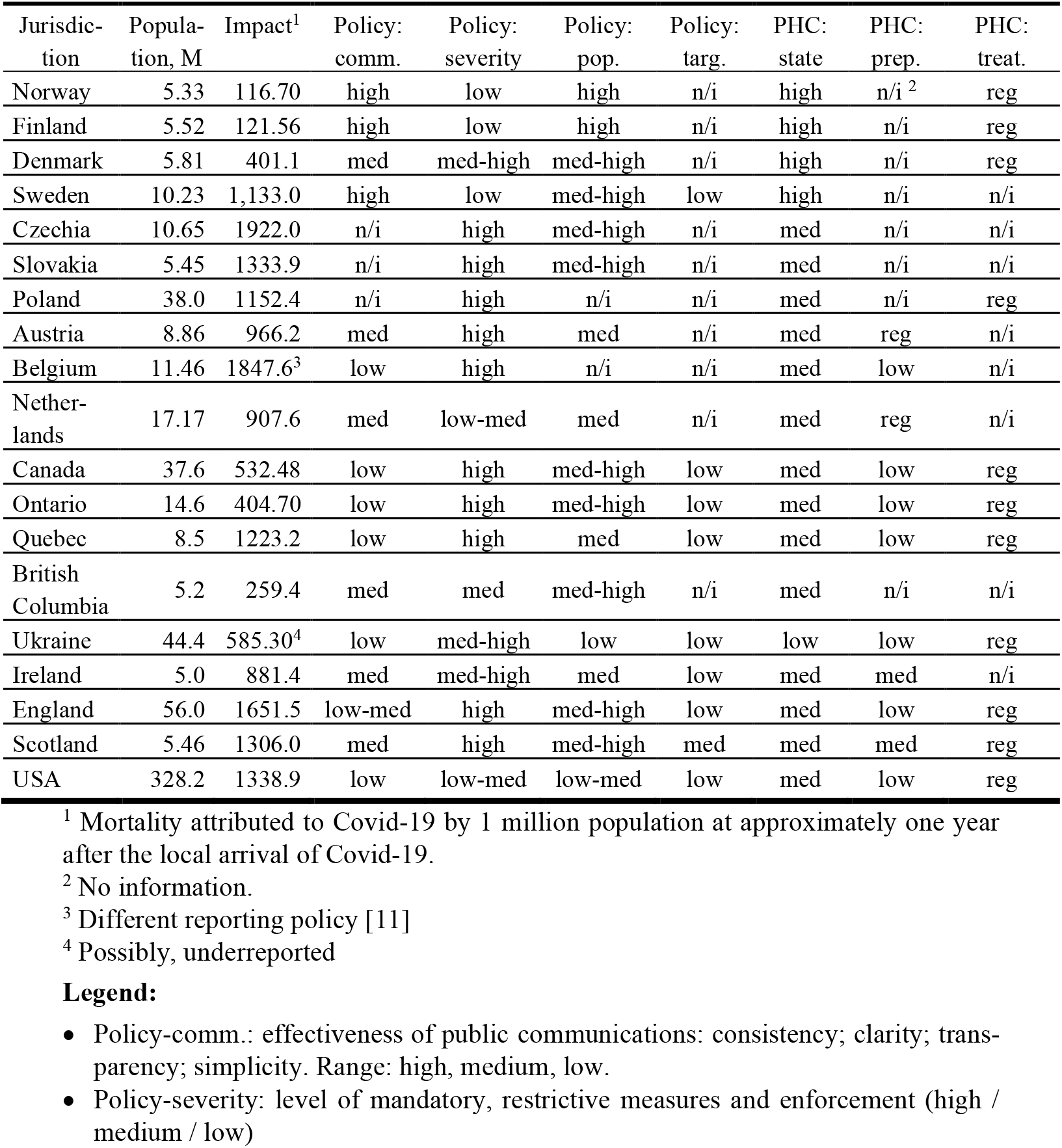
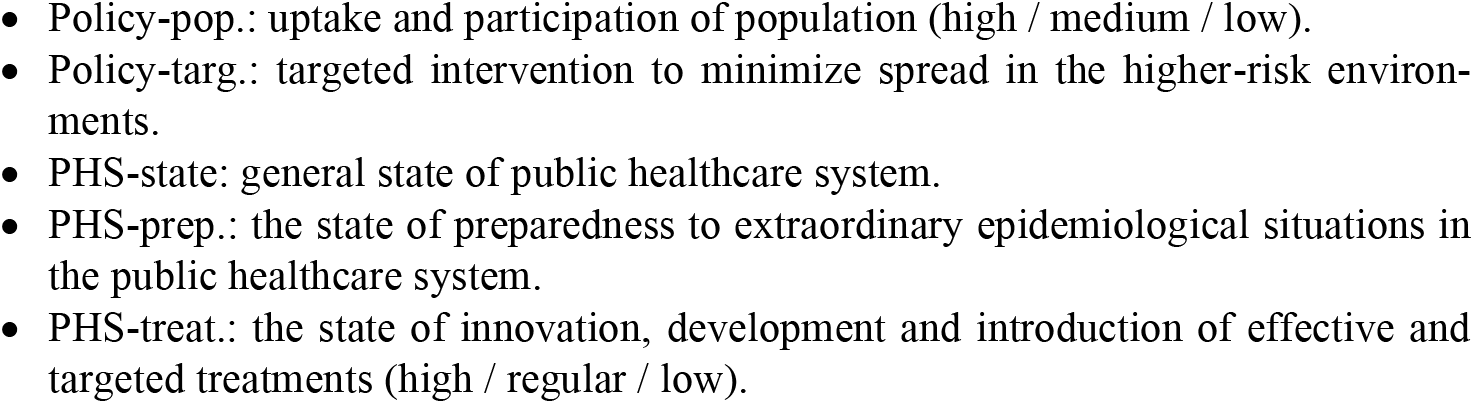
National jurisdictions pVCE dataset: policy factors

| Jurisdiction | Population, M | Impact<sup>1</sup> | Policy: comm. | Policy: severity | Policy: pop. | Policy: targ. | PHC: state | PHC: prep. | PHC: treat. |
| --- | --- | --- | --- | --- | --- | --- | --- | --- | --- |
| Norway | 5.33 | 116.70 | high | low | high | n/i | high | n/i <sup>2</sup> | reg |
| Finland | 5.52 | 121.56 | high | low | high | n/i | high | n/i | reg |
| Denmark | 5.81 | 401.1 | med | med-high | med-high | n/i | high | n/i | reg |
| Sweden | 10.23 | 1,133.0 | high | low | med-high | low | high | n/i | n/i |
| Czechia | 10.65 | 1922.0 | n/i | high | med-high | n/i | med | n/i | n/i |
| Slovakia | 5.45 | 1333.9 | n/i | high | med-high | n/i | med | n/i | n/i |
| Poland | 38.0 | 1152.4 | n/i | high | n/i | n/i | med | n/i | reg |
| Austria | 8.86 | 966.2 | med | high | med | n/i | med | reg | n/i |
| Belgium | 11.46 | 1847.6<sup>3</sup> | low | high | n/i | n/i | med | low | n/i |
| Netherlands | 17.17 | 907.6 | med | low-med | med | n/i | med | reg | n/i |
| Canada | 37.6 | 532.48 | low | high | med-high | low | med | low | reg |
| Ontario | 14.6 | 404.70 | low | high | med-high | low | med | low | reg |
| Quebec | 8.5 | 1223.2 | low | high | med | low | med | low | reg |
| British Columbia | 5.2 | 259.4 | med | med | med-high | n/i | med | n/i | n/i |
| Ukraine | 44.4 | 585.30<sup>4</sup> | low | med-high | low | low | low | low | reg |
| Ireland | 5.0 | 881.4 | med | med-high | med | low | med | med | n/i |
| England | 56.0 | 1651.5 | low-med | high | med-high | low | med | low | reg |
| Scotland | 5.46 | 1306.0 | med | high | med-high | med | med | med | reg |
| USA | 328.2 | 1338.9 | low | low-med | low-med | low | med | low | reg |

<sup>1</sup> Mortality attributed to Covid-19 by 1 million population at approximately one year after the local arrival of Covid-19.
<sup>2</sup> No information.
<sup>3</sup> Different reporting policy [11]
<sup>4</sup> Possibly, underreported

**Table 2.**
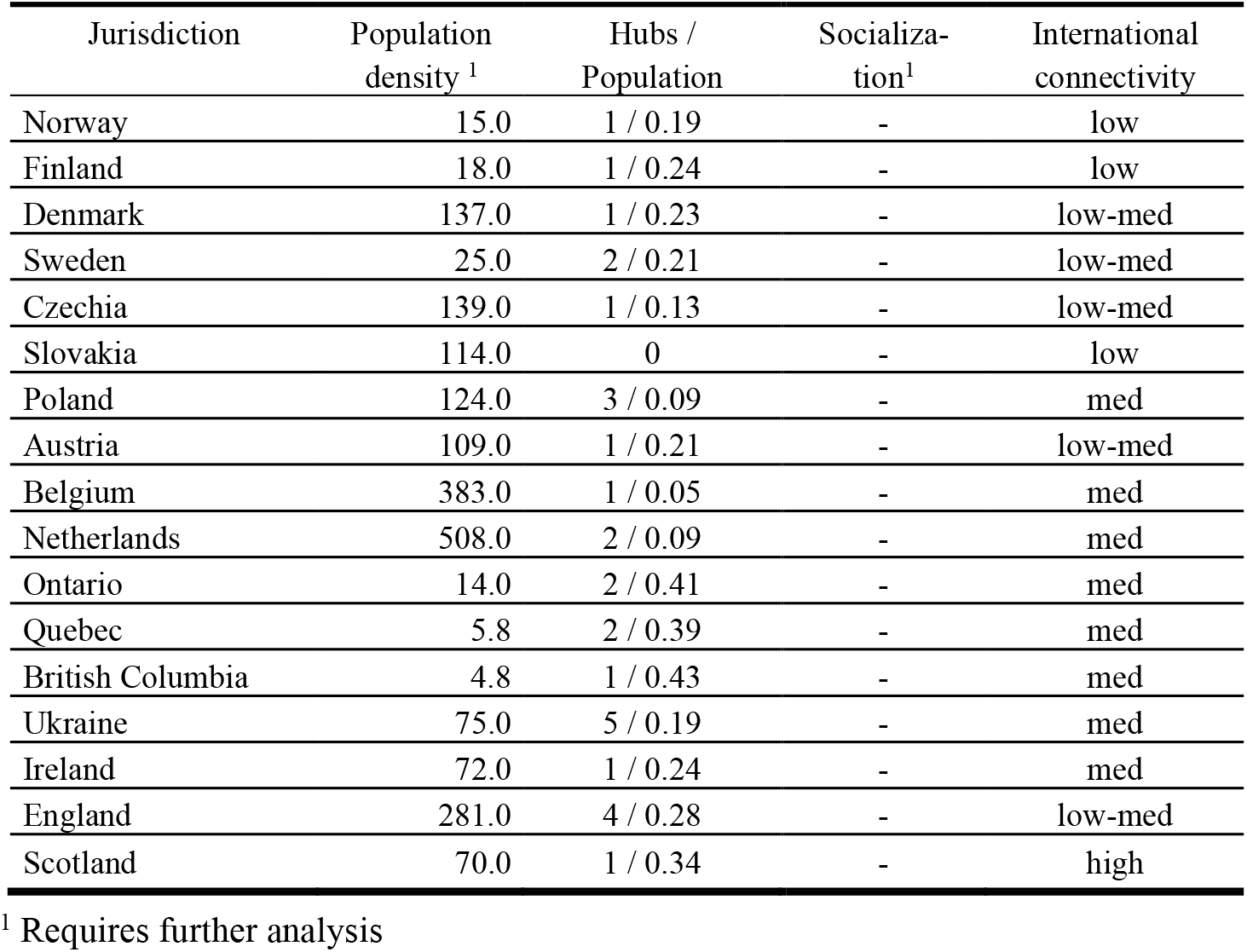
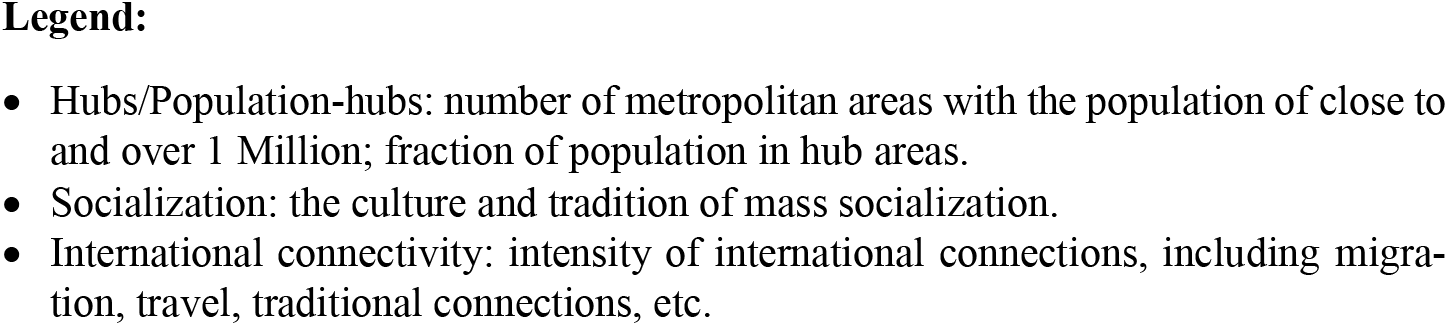
National jurisdictions pVCE dataset: social factors

| Jurisdiction | Population density<sup>1</sup> | Hubs / Population | Socialization<sup>1</sup> | International connectivity |
| --- | --- | --- | --- | --- |
| Norway | 15.0 | 1 / 0.19 | - | low |
| Finland | 18.0 | 1 / 0.24 | - | low |
| Denmark | 137.0 | 1 / 0.23 | - | low-med |
| Sweden | 25.0 | 2 / 0.21 | - | low-med |
| Czechia | 139.0 | 1 / 0.13 | - | low-med |
| Slovakia | 114.0 | 0 | - | low |
| Poland | 124.0 | 3 / 0.09 | - | med |
| Austria | 109.0 | 1 / 0.21 | - | low-med |
| Belgium | 383.0 | 1 / 0.05 | - | med |
| Netherlands | 508.0 | 2 / 0.09 | - | med |
| Ontario | 14.0 | 2 / 0.41 | - | med |
| Quebec | 5.8 | 2 / 0.39 | - | med |
| British Columbia | 4.8 | 1 / 0.43 | - | med |
| Ukraine | 75.0 | 5 / 0.19 | - | med |
| Ireland | 72.0 | 1 / 0.24 | - | med |
| England | 281.0 | 4 / 0.28 | - | low-med |
| Scotland | 70.0 | 1 / 0.34 | - | high |

<sup>1</sup> Requires further analysis
- Hubs/Population-hubs: number of metropolitan areas with the population of close to and over 1 Million; fraction of population in hub areas. - Socialization: the culture and tradition of mass socialization. - International connectivity: intensity of international connections, including migration, travel, traditional connections, etc.

#### Notes

##### 1. Policy, severity

Ontario, Quebec, Austria, Poland, Belgium, Slovakia, England: three hard lockdowns; Ukraine, Czechia, Belgium, Ireland, Scotland, Denmark: two hard lockdowns; Netherlands: one lockdown. Norway: one short lockdown (two weeks).

##### 2. Policy, targeted intervention

Senior residences and retirement homes; other mass residences; workplaces; schools and colleges

##### 3. PHS preparedness

Shortage of medical supplies, first response, emergency, critical care capacity, strained critical care facilities, and similar.

## 3 Analysis

As can be observed in Table 1, no significant correlation between the severity of quarantine policy and the epidemiological impact can be observed. The observation is even more apparent with the subset of northern state cases: Ontario, Quebec, British Columbia, Scotland and the four Scandinavian countries. It can be confirmed by formal calculation of the Pierson coefficient between the observed impact and a factor of severity of the quarantine policy determined based on the number and duration of hard quarantines. As shown further in this section, the result appears to be midrange positive, i.e. the severity of quarantine weakly correlated with higher epidemiological impact, quite contrary to intuitive expectation of a desired outcome, that is, a reduction in epidemiological impact.

The dataset contains interesting representative samples of different approaches in epidemiological policies aimed at controlling the infection, though any conclusions need to be made with caution given the small number of representatives. There are policies aimed at effective integrated management with the objective of minimization of both epidemiological impact, and disruption to the society (IEM): Norway, Finland, Sweden, British Columbia. On the opposite of the spectrum there is strict control group, with multiple strict quarantines (SQ) represented by Austria, Belgium, Ontario, Quebec and other jurisdictions. There seems to be a group employing quarantines on as-necessary basic (NQ), Denmark, Scotland and others. There is also an example of a “default scenario” with minimal effectiveness of the policy (ME), Ukraine. However, it did not contain instances of another strategy of “total suppression” applied successfully in a number of countries (Australia, New Zealand, Taiwan and others) that encompasses aggressive tracking of individual cases and strict but limited in duration and geographical span “blitz” quarantines [12] to suppress outbreaks identified in the early stage with intelligent and ubiquitous testing. It can be viewed as a variant of IEM however a detailed analysis and discussion of this approach will be provided elsewhere.

Certainly, policy factors undoubtedly important as they are, are not the only ones that can impact the course of the epidemics that would not be possible to analyze or even discuss here. More detailed analysis of this factors and their influence is an interesting objective for a future study.

To understand the association between policy factors and the observed epidemiological impact a two-factor analysis was performed. A numerical factor indicating the level of severity of the quarantine policy, 0 < *P*_*qr*_ < 1, was introduced for jurisdictions in the study as outlined earlier. We also attempted to define a factor of effectiveness of epidemiological response *P*_*eff*_, with the same numerical range, determined as follows:

1. General level of quality of public healthcare system taken as a base (0.2 – 0.8, poor to excellent).
2. Points added or subtracted based on the following factors: quality of monitoring and timeliness of action; clarity and consistency of communication; effectiveness of targeted intervention (e.g., senior residences, workplaces, gatherings, etc.); quality of epidemiological management in the public healthcare (e.g., resources, capacity, introduction of effective treatments, etc.)

Other factors were assumed to have lower influence on the epidemiological outcome. Thus, equalization of factors other than policy was an essential consideration in determination of selection criteria in Section 2.1. The resulting table of factors can be found in the Appendix (Table A1).

Calculated values of the correlation coefficient for factors *P*_*qr*_, *P*_*eff*_ and the recorded epidemiological outcomes in the jurisdictions in pVCE dataset were, respectively, *0*.*451* (positive) and *-0*.*497* (negative) i.e. higher severity of the quarantine policy reflected by factor *P*_*qr*_ was correlated with higher epidemiological outcome, whereas higher effectiveness of the policy described by factor *P*_*eff*_ appeared to have a midrange positive correlation with lower outcomes.

The relationship of *P*_*qr*_, *P*_*eff*_ and the recorded impact is illustrated in Figure 1 as a scatter plot of policy factors with minimum squared error linear trends.

**Fig. 1.**
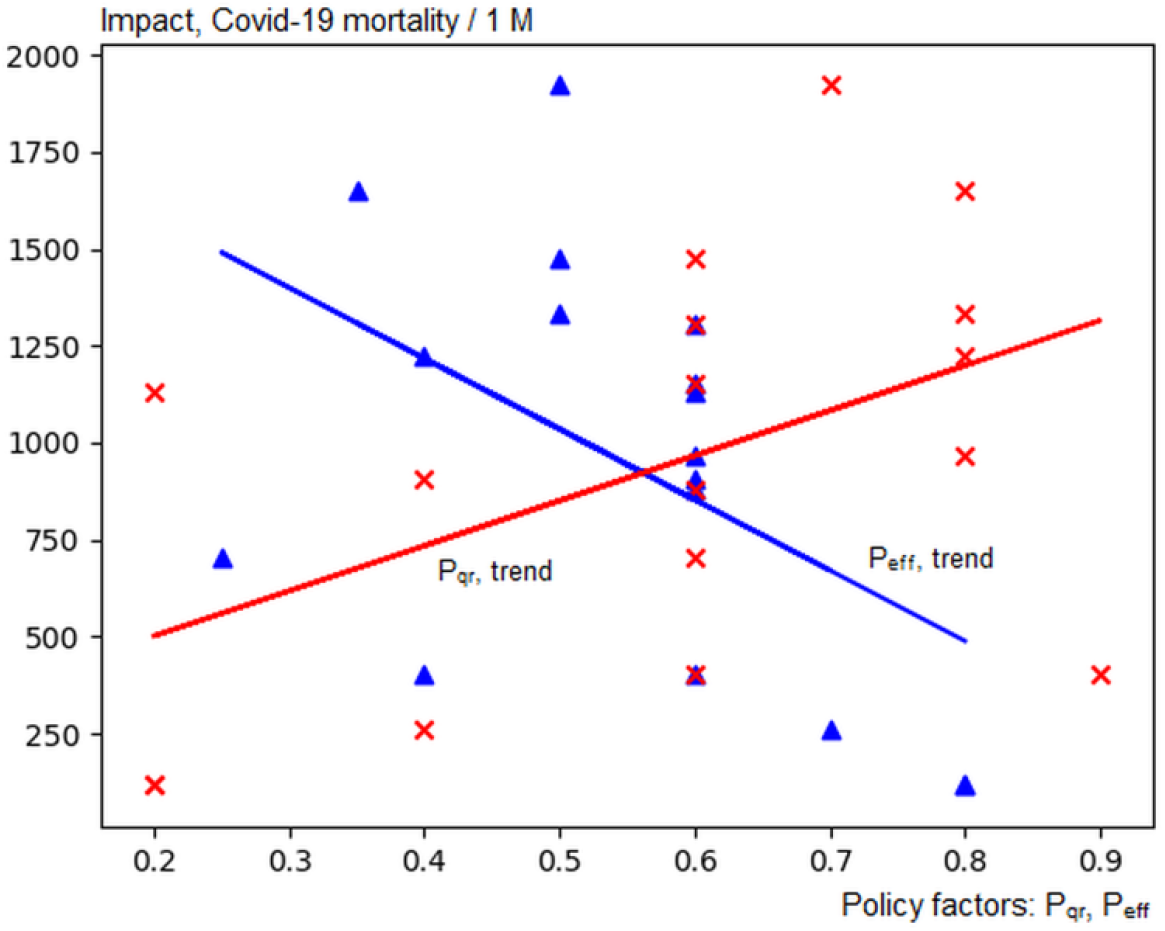
Epidemiological impact vs. policy effectiveness (*P*_*eff*_, blue) and quarantine policy severity (*P*_*qr*_, red) with min. squared error linear trends.

## 4 Discussion

Positive (and ostensibly, contrary to the intent of minimization of the epidemiological impact) correlation between the severity of quarantine policy and the observed epidemiological impact is not necessarily surprising, given the frequently used rationale that strict quarantines prevent even more severe outcomes in the scenarios with a rapid onset of the infection (such as developing “waves” of infection). When a strict lockdown is introduced in a situation with rapidly increasing case counts and raising epidemiological impact, the association, and correlation becomes that of the reaction, cause and effect, a new wave of cases (likely with a higher epidemiological impact) to a strict quarantine.

Two observations can be made in this regard. First, it can be observed that entering a scenario with rapid and broad, community-wide spread of the infection itself can be the result of shortcomings in development and implementation of effective epidemiological policy, including: current and accurate monitoring of the situation; open and effective communications with the population; effective targeted intervention where and whenever needed; and not in the least, management, preparation, adjustment and continuous improvements in the public healthcare system, itself a broad topic for a separate study. If and when all of these components of an effective policy are brought together in an integrated response, one would need to estimate the potential for “explosive” scenarios and design further action plans. That state however, does not appear to have been attained in the majority of considered cases.

And in such cases, an unintended outcome can be that the pattern of strict and stricter quarantines seen as, and becoming the de-facto and default solution taken as an automatic reaction to any significant increase in cases without consideration of other options. If extended over longer periods these policies can be taxing and disruptive for the society, and possibly not sustainable in the longer run (Ontario, with strict quarantine continuing, with a brief interruption, from end of December, 2020 to early June 2021, i.e. close to five months at the time of writing). It is not clear how far such long quarantines can be extended without causing significant disruption in the fabric of the society and a backlash or indifference in the population, yet locking into the pattern “any spike in cases leads to broad and strict quarantine” may cause reluctance to relax restrictions at the appropriate time due to desire to avoid the risk of the resurgence in the absence of other effective instruments of controlling the infection, locking the society in the less productive from the perspective of overall minimization of impact trend that was observed in the previous section.

In the authors view, an effective epidemiological response to Covid-19 and future pandemics can be based on four important and necessary cornerstones integrated in a single effective policy:

- Current, accurate, detailed and comprehensive monitoring of the situation.
- Clear, open, honest and active communications with the population, maintaining both trust and engagement, with the focus on voluntary participation and compliance.
- Effective targeted intervention in the areas and environments of increased or increasing epidemiological risk.
- Effective management, adjustment and improvement of the epidemiological capacity in the public health care in all relevant areas, including capacity, resources, connection to research for effective treatments and other.

Following this strategy to a comprehensive implementation would provide a broad range of effective tools and methods of controlling epidemiological situation [13], including when and if necessary, targeted quarantines as one of the instruments in the toolkit, however maybe not the only one and the default one.

In the end the question that in our view needs to be studied and answered is, what are the objectives of the policy? They need to be defined, discussed and made public before the policy is put in place to know how effective it is (or perhaps, not). With the integrated effective management approach, the objectives to minimize the impact of the epidemics and disruption to the society are set simultaneously, emphasizing methods and instruments that allow to control the epidemics effectively while being compatible with normal, or near-normal functioning of the society. Progress in both objectives can be verified quantitatively and corrective measures designed and implemented if and as necessary.

With the reactive approach, on the other hand, the question of effectiveness becomes less clear as there is no clear quantitative benchmark against which the performance of the policy can be evaluated. How effective were restrictive quarantines? Did they achieve their objectives? These questions may not be straightforward to answer without introducing hypothetical scenarios with unknown accuracy (in one case a model prediction of up to 18,000 daily cases [14] was used in Ontario as a rationale for further restrictive measures; it never came to pass, even by a remote proximity. Did it speak for the success of the policy, or the accuracy of the models on which it was based? There is no easy answer to this question and it is not clear what has been learned and achieved). As the results of this work indicate, even the primary objective of minimizing the epidemiological impact may not be achievable readily and reliably with such a strategy without extreme restrictions with a high cost to the society.

## Data Availability

The data is available upon request to the corresponding author.

https://www.google.com/covid19-map

https://www.worldometers.info/world-population/

https://en.wikipedia.org/wiki/Template:COVID-19_pandemic_data

## Appendix

**Table A1.** Policy factors, pVCE dataset

| Jurisdiction | Quarantine severity,<br>$P_{qr}$ | Policy effectiveness,<br>$P_{eff}$ |
| --- | --- | --- |
| Norway | 0.2 | 0.8 |
| Finland | 0.2 | 0.8 |
| Denmark | 0.6 | 0.6 |
| Sweden | 0.2 | 0.6 |
| Czechia | 0.7 | 0.5 |
| Slovakia | 0.8 | 0.5 |
| Poland | 0.6 | 0.6 |
| Austria | 0.8 | 0.6 |
| Belgium | 0.6 | 0.5 |
| Netherlands | 0.4 | 0.6 |
| Ontario | 0.9 | 0.4 |
| Quebec | 0.8 | 0.4 |
| British Columbia | 0.4 | 0.7 |
| Ukraine | 0.6 | 0.25 |
| Ireland | 0.6 | 0.6 |
| England | 0.8 | 0.35 |
| Scotland | 0.6 | 0.6 |

## Notes

### Competing Interest Statement

The authors have declared no competing interest.

### Funding Statement

This research received no specific funding

### Author Declarations

Exemption: only open data available without registration used in the work.

